# Development and Validation of Early Warning Score Systems for COVID-19 Patients

**DOI:** 10.1101/2020.11.04.20225904

**Authors:** Alexey Youssef, Samaneh Kouchaki, Farah Shamout, Jacob Armstrong, Rasheed El-Bouri, Thomas Taylor, Drew Birrenkott, Baptiste Vasey, Andrew Soltan, Tingting Zhu, David A Clifton, David W Eyre

## Abstract

COVID-19 is a major, urgent, and ongoing threat to global health. Globally more than 24 million have been infected and the disease has claimed more than a million lives as of October 2020. Predicting which patients will need respiratory support is important to guiding individual patient treatment and also to ensuring sufficient resources are available. We evaluated the ability of six common Early Warning Scores (EWS) to identify respiratory deterioration defined as the need for advanced respiratory support (high-flow nasal oxygen, continuous positive airways pressure, non-invasive ventilation, intubation) within a prediction window of 24 hours. We show these scores perform sub-optimally at this specific task. Therefore, we develop an alternative Early Warning Score based on a Gradient Boosting Trees (GBT) algorithm that is able to predict deterioration within the next 24 hours with high AUROC 94% and an accuracy, sensitivity and specificity of 70%, 96%, 70%, respectively. Our GBT model outperformed the best EWS (LDTEWS:NEWS), increasing the AUROC by 14%. Our GBT model makes the prediction based on the current and baseline measures of routinely available vital signs and blood tests.

## 1. Introduction

COVID-19 is a major, urgent, and ongoing threat to global health. The disease has infected millions across the globe and caused more than 1 million deaths due to direct infections as of 9^th^ October [1], causing a surge in demand on healthcare services. This has created a significant strain on hospital resources globally, especially on intensive care units (ICUs) and respiratory support equipment such as invasive and non-invasive ventilators, on which the survival of severely ill patients depends. In such conditions, a tool to predict deterioration of patients in advance is very valuable to best allocate hospital resources. Moreover, such a tool is helpful for individual patient care as it would ensure that patients are placed in the correct environment to meet their needs, e.g. transferred to ICU before substantial deterioration. Given the limited resources there is a significant need to prioritise the right patients so the resource is available for those who need it.

Deterioration prediction tools have traditionally existed in the form of Early Warning Score (EWS) systems or “Physiological Track and Trigger Systems”, which track physiological variables and alert for assistance when the variables surpass a predefined threshold [2]. The first scores were developed in 1997 by Morgan and colleagues [2] as a tool to ensure the presence of trained medical staff by the bedside of deteriorating patients exhibiting physiological derangement. Thus, these scores were developed to detect deterioration based on the premise that physiological instability precedes critical clinical deterioration [3]. Early EWS systems use an aggregate weighted scoring of vital signs, roughly the same approach in use by the plethora of available scores today [3]. Although EWS are not intended to predict adverse events, but rather to alert for deterioration that may precede adverse events, different scores have been developed, adjusted and validated with this goal, e.g. predicting unexpected ICU admission, in-hospital mortality, and cardiac arrest, among others [2].

EWS systems that are currently used in practice are based on routinely measured physiological variables, including vital signs, consciousness level (AVPU), oxygen support, and different laboratory markers [4, 3, 5, 6, 7]. Common examples of these tools include the National Early Warning Score (NEWS) [7], Centile-Based Early Warning Score (CEWS) [3], Manual Centile-Based Early Warning Score (MCEWS) [4], Age-Based Early Warning Score (AEWS) [8], Laboratory Decision Tree Early Warning Score (LDTEWS) [9], and LDTEWS:NEWS [5] (which combines both NEWS and LDTEWS), among others. The scores evaluate physiological parameters (NEWS, CEWS, MCEWS and AEWS), laboratory parameters (LDTEWS) and a combination of physiological and laboratory parameters (LDTEWS:NEWS) (Supplementary A: Table S6) [7, 3, 4, 8, 9, 5]. They have also been validated for different outcomes, including ICU admission, mortality, and cardiac arrest, usually within 24 hours from the time of collecting the vital-sign measurements or obtaining the laboratory test results (Supplementary A: Table S6).

It is currently unclear how valuable EWS systems would be in predicting deterioration amongst COVID-19 patients, since these scores have been developed and validated for all causes of deterioration in a pre-pandemic in-patient cohort. By contrast, deterioration in the COVID-19 inpatient cohort more commonly manifests through hypoxic respiratory failure [10]. Therefore, EWS tools built to detect deterioration in the general patient population may be less effective at predicting deterioration in COVID-19 patients. This is reflected in structural differences in the EWS calculation algorithms. For example, NEWS considers a binary variable for oxygen support (room air vs. oxygen support) while the rest of the variables are either continuous or categorical [7]. Therefore, NEWS is not equipped to capture the variability in the different oxygen support levels (low-flow nasal cannulae to invasive ventilation), which can be a strong proxy for severity of COVID-19 respiratory distress. However, despite the limitations, these tools are in routine use in many hospital managing patients with COVID-19.

To support the management of hospitalised COVID-19 patients, different EWS have been used or developed to predict patient deterioration. Researchers have also tried to validate the existing EWS on COVID-19 patients [11, 12, 13, 14, 15]. For example, the performance of NEWS has been tested on hospitalised, community (off-label), and care home COVID-19 patients (off-label) [13, 12, 15]. Moreover, Meylan et al. and Carr et al. have explored adjustments to the score (NEWS2) to adapt it for COVID-19 patients specifically [16, 17]. Carr and colleagues validated the ability of the NEWS2 score to identify severe COVID-19 infections (defined as ICU admission or in-hospital mortality). The study reported an initial performance of 0.628 area under the receiving operator characteristic curve (AUROC) and a performance of 0.753 AUROC after including five additional predictive features: age, c-reactive protein (CRP), neutrophil count, estimated Glomerular Filtration Rate (GFR), and Albumin [16]. The Royal College of Physicians has endorsed the use of (NEWS2) to predict deterioration of COVID-19 patients [18]. NEWS2 has been evaluated by Meylan et al. and by Carr et al. [16, 17]; however, many scores have not yet been evaluated appropriately, including NEWS, CEWS, MCEWS, AEWS, LDTEWS, and LDTEWS:NEWS. Therefore, it is critical to evaluate the performance of different available EWS on a COVID-19 inpatient population, to inform clinical practice during the global pandemic.

In addition to EWS, the medical community has developed Machine Learning (ML) tools that can be used to improve the management of COVID-19 patients. Multiple models have been developed to use CT images [19] and routinely collected clinical data (routine blood tests, venous/arterial gasses, and vital signs) [20] to support the diagnosis of COVID-19. These models report a performance (AUROC) that ranges from 0.81 to nearly 1 [19]. Other prognostic machine learning models have been developed, predicting mortality, length of hospital stay, and progression to a severe or critical state [21, 22, 23, 24, 25, 26]. However, some of these models have methodological limitations like sampling bias; for example, some of the prognostic models that predicted mortality had a mortality rate that ranged from 8% to 56%, a rate that is higher than that in the general COVID-19 inpatient cohort [19, 21, 27, 24, 25].

The reported performance of EWS and ML models in the literature is high. However, a recent systematic review by Wynants and colleagues has critically appraised these COVID-19 diagnosis and prognosis prediction models [19]. The study highlights that these models lack validation and calibration and are at high risk of bias (evaluated through the PROBAST tool [28]) in multiple domains: participants, predictors, outcome domains, among others. According to Wynants and colleagues, the high risk of bias suggests that the predictive performance of the scores is likely lower than reported, and that the generated predictions are not reliable. Although it is critical to have timely access to predictive tools that can optimise patient triage and resource utilisation within hospitals, such tools need to be appropriately designed and sufficiently validated before adoption into clinical care. Improperly validated EWS systems and predictive models are of limited clinical benefit in pandemic settings and their risk of harm can potentially outweigh the promised benefit [19, 29, 30].

In response to the aforementioned clinical and research need to validate EWS in COVID-19 patients, our work has three main contributions: (i) we evaluate the performance of existing EWS systems (Supplementary A: Table S6) that may be currently used in practice to predict clinical deterioration in COVID-19 patients, (ii) we develop and validate machine learning models to predict deterioration in advance for COVID-19 patients, and (iii) we compare between machine learning methods and traditional EWS systems.

## 2. Results

### 2.1. Patient cohort and features characteristics

Our study is retrospective, using data extracted from electronic health records (EHR). The dataset contains routinely collected observations from concluded hospital admissions from four hospitals within the Oxford University Hospitals (OUH) NHS Foundation Trust. OUH consists of 4 teaching hospitals in Oxfordshire, UK, serving a population of 600,000 and providing tertiary referral services to the surrounding region. Data were obtained between March and July 2020.

Our dataset included 15,686 sets of observations from 472 admissions in 472 unique patients. The dataset included 137 respiratory deterioration events (observing each patient until discharge or their first deterioration). The average age was 68 ± 16 (mean ± std) and 47% (221/472 patients) of the dataset were females. The mean and interquartile ranges (IQR) of the blood, blood gas, and vital sign parameters are in the supplementary materials (Supplementary A: Tables S2, S3, and S4).

### 2.2. Performance evaluation

#### 2.2.1. Outcome definition

We defined respiratory deterioration as the need for advanced respiratory support (high-flow nasal oxygen [HFN0], continuous positive airways pressure [CPAP], non-invasive ventilation [NIV], intubation) or ICU admission within a prediction window of 24 hours. It should be noted, however, that hypoxic respiratory failure is not the only process through which COVID-19 patients deteriorate as some patients deteriorate through a process of shock due to venous thromboembolism or super-added sepsis. Such events may also lead to ICU admission or increased oxygen requirements and so still be captured by our model.

#### 2.2.2. Performance of the EWS systems

Table 2 outlines the performance of the EWS systems. NEWS, MCEWS, CEWS, AEWS, LDTEWS:NEWS, and LDTEWS achieved an AUROC of 79%, 78%, 63%, 68%, 80%, 62%, respectively. The best performing scores were NEWS and LDTEWS:NEWS.

We evaluated the performance of the recommended (original) thresholds for the different Early Warning Scores. The default thresholds are 5, 4, 4, 0.27, 0.33 for NEWS, CEWS, MCEWS, LDTEWS:NEWS, and LDTEWS, respectively. AEWS does not have a recommended threshold, therefore we have excluded it from the evaluation of the recommended thresholds. The NEWS score had the most balanced Sensitivity and Specificity (66% and 75% respectively). NEWS and LDTEWS achieved the lowest accuracy (75% and 73%) with a sensitivity and specificity of 41% and 74% for LDTEWS. CEWS achieved the highest accuracy (91%) but with a sensitivity of 23% and specificity of 91% (Table 2).

We optimised the thresholds for each score to maximise accuracy as outlined in the Methods Section. Optimised EWS thresholds yielded more balanced performance. LDTEWS:NEWS was the overall best performing score with an accuracy, sensitivity, and specificity of 67%, 77% and 67%, respectively. NEWS, MCEWS, CEWS, and AEWS achieved high accuracy (62%, 61%, 64%, 60%, respectively). The worst performing score was LDTEWS with an accuracy of 52% and AUROC of 62%. The accuracy-optimised thresholds for all scores differed from the recommended values (Table 2).

The performance of the EWS in COVID-19 patients was significantly lower than that previously reported in non-COVID patients. The Royal College of Physicians [7] reported a performance of (AUROC = 89%) for NEWS compared to (AUROC = 79%) in our dataset. Watkinson and colleagues [4] reported a performance of (AUROC = 86.8% and AUROC = 80.8%) for MCEWS and CEWS, respectively. This compares to AUROC values of 78% and 63% for MCEWS and CEWS in our dataset. Shamout and colleagues [8] reported that AEWS achieved an AUROC of 83.8%, while AEWS achieved a performance of 68% on COVID patients in our dataset. Redfern and colleagues reported an AUROC of 90.1%-91.6% for LDTEWS:NEWS. In COVID patients, the AUROC for LDTEWS:NEWS was 80%. The worst performing score in our study was LDTEWS (AUROC of 62%). The score was developed by Jarvis and colleagues [9] with a reported AUROC that ranges between 75-80% in discriminating in-hospital mortality among the general in-hospital patient cohort. This indicates that while the predictors used in LDTEWS (HGB, Alb, Na, k, Cr, Ur, WBC) are useful to discriminate in-hospital mortality in non-COVID, they are less useful in predicting respiratory deterioration in COVID patients (Supplementary A: Table S6).

### 2.2.3. Performance of the machine learning models

We evaluated the performance of three machine learning models (Gradient Boosted Trees [GBT], Random Forest [RF], Logistic Regression [LR]) on the training data using an internal 5-fold cross validation. We evaluated the performance of the ML models on multiple feature sets as outlined in the feature sets subsection of the Methods and Table 1 (F1-F11). The GBT model outperformed the other models on the different features sets in our training dataset. Therefore, we made a design choice to use only the GBT model when evaluating the performance on the different feature sets in the test data. The highest AUROC was achieved using the F1 (AUROC of 83%), F7 (AUROC of 93%), F8 (AUROC of 86%), F9 (AUROC of 94%), and F11 (AUROC of 93%) feature sets. The lowest AUROC was observed in the F2 (AUROC of 72%), F4 (AUROC of 77%), F5 (AUROC of 69%), and F6 (AUROC of 78%) feature sets.

**Table 1:**
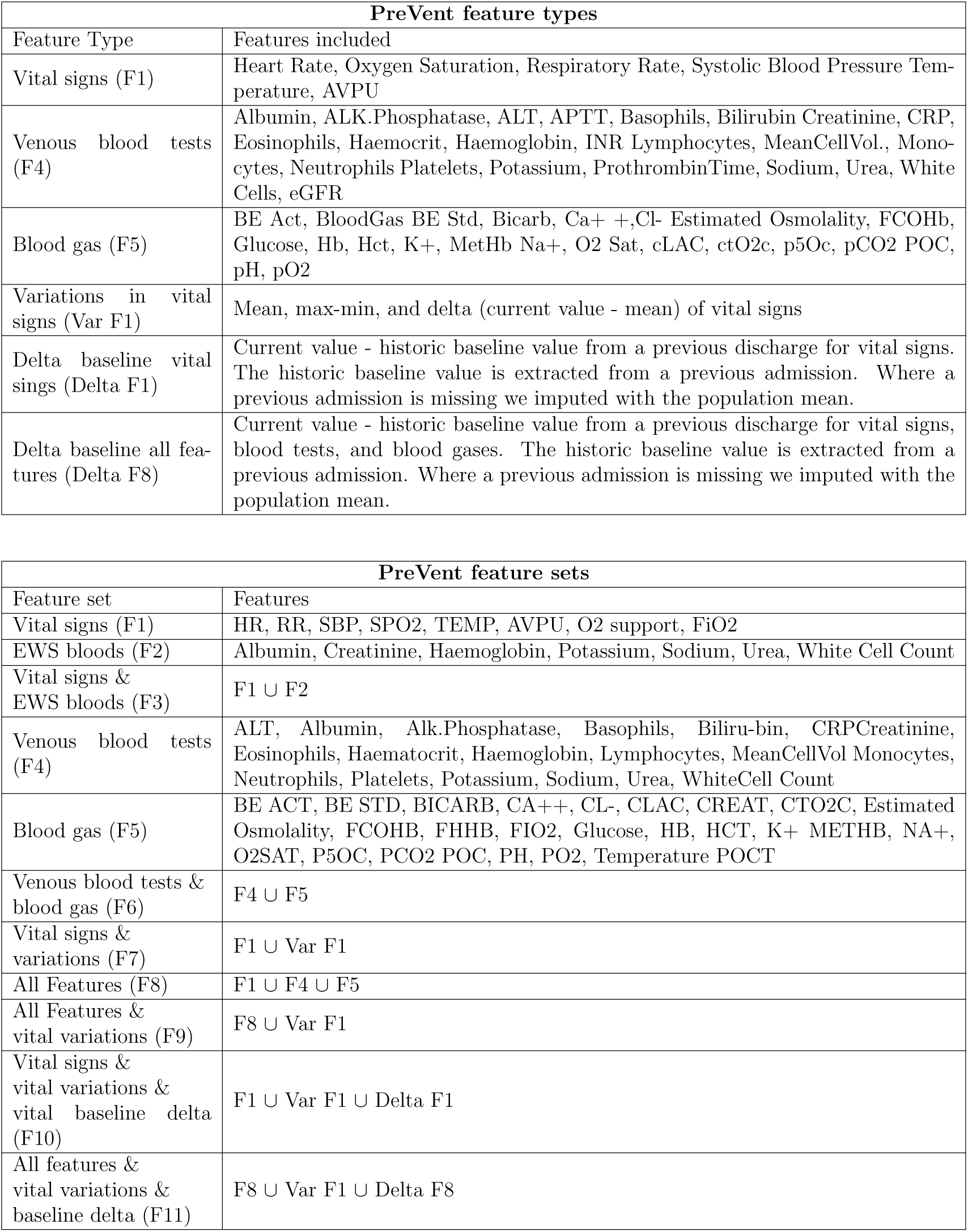
The different feature types and feature sets in our study.

We compared the performance of the EWS systems and machine learning models to predict COVID-19 patient deterioration in three main feature sets: F1-F3 (Table 1). In each of the three feature sets, the machine learning model outperformed the Early Warning Scores. For the F1 feature set, we can compare the performance of NEWS (AUROC = 79%), MCEWS (AUROC = 78%), CEWS (AUROC = 63%), and AEWS (AUROC = 68%) with the performance of GBT (AUROC = 83%). For the F2 feature set, we can compare the performance of LDTEWS (AUROC = 62%) with the performance of GBT (AUROC = 72%). For the F3 set, we can compare the performance of LDTEWS:NEWS (AUROC = 80%) with the performance of GBT (AUROC = 85%).

The overall best performing algorithm for machine learning models was the GBT model on the F9 feature set (AUROC = 94%). Given the imbalanced nature of our dataset, we have decided to tune the probability-class conversion threshold for the GBT model to create the best performing machine learning model. We decided to optimise the threshold to maximise accuracy. We identified the threshold that maximises the accuracy of the GBT model on the training set and measured the performance on the test set. The identified threshold was 0.19. The optimised GBT model achieved an accuracy, sensitivity, and specificity of 70%, 96%, and 70%, respectively. The most and least important features are outlined in Table 3. Out of the ten most important features (FiO2, min-max SBP, CRP, max-min HR, PO2, mean cell volume, arterial blood calcium, max-min RR, CtO2C, temp), four belonged to the F7 (vital signs & variability) feature set, three belonged to the F5 feature set (arterial blood tests), and two belonged to the F4 feature set (venous blood tests). The most important feature was FiO2. Delta is a measure of variability of a specific variable, it is calculated as (current value - the mean in the last 24 hours). The most important vital signs were heart rate, respiratory rate, temperature, and blood oxygen saturation (SpO2).

**Table 2:** This table highlights the performance of the Early Warning Scores. Section A of the table describes the performance of the original thresholds for the Early Warning Scores. NEWS has two recommended thresholds (5 and 7) and LDTEWS:NEWS also has two recommended thresholds (0.27 and 0.36). We measured the performance of 5 and 0.27 for NEWS and LDTEWS:NEWS respectively as those are the most commonly used thresholds. Section B outlines the performance of the accuracy-optimised thresholds.

| The performance of the original thresholds (Section A) |  |  |  |  |  |  |
| --- | --- | --- | --- | --- | --- | --- |
| Score | Threshold | Acc | Sen | Sps | Prs | AUROC |
| NEWS | 5 | 0.75 (0.75-0.75) | 0.66 (0.65-0.68) | 0.75 (0.75-0.75) | 0.02 (0.02-0.02) | 0.79 (0.78-0.79) |
| CEWS | 4 | 0.91 (0.91-0.91) | 0.23 (0.21-0.24) | 0.91 (0.91-0.91) | 0.02 (0.02-0.02) | 0.63 (0.61-0.64) |
| MCEWS | 4 | 0.78 (0.78-0.78) | 0.61 (0.59-0.62) | 0.78 (0.78-0.78) | 0.02 (0.02-0.02) | 0.78 (0.77-0.79) |
| LDTEWS | 0.33 | 0.73 (0.73-0.73) | 0.41 (0.40-0.43) | 0.74 (0.73-0.74) | 0.01 (0.01-0.01) | 0.62 (0.61-0.63) |
| LDTEWS:NEWS | 0.27 | 0.84 (0.84-0.84) | 0.52 (0.50-0.53) | 0.84 (0.84-0.85) | 0.03 (0.03-0.03) | 0.80 (0.79-0.80) |

| The performance of the accuracy-optimised thresholds (Section B) |  |  |  |  |  |  |
| --- | --- | --- | --- | --- | --- | --- |
| Score | Threshold | Acc | Sen | Sps | Prs | AUROC |
| NEWS | 4 | 0.62 (0.60-0.64) | 0.77 (0.75-0.79) | 0.62 (0.60-0.64) | 0.02 (0.02-0.02) | 0.79 (0.78-0.79) |
| CEWS | 2 | 0.64 (0.62-0.65) | 0.55 (0.52-0.57) | 0.64 (0.62-0.65) | 0.01 (0.01-0.01) | 0.63 (0.61-0.64) |
| MCEWS | 3 | 0.61 (0.59-0.63) | 0.76 (0.74-0.79) | 0.61 (0.59-0.62) | 0.02 (0.02-0.02) | 0.78 (0.77-0.79) |
| AEWS | 4 | 0.60 (0.59-0.62) | 0.66 (0.64-0.68) | 0.60 (0.59-0.62) | 0.01 (0.01-0.01) | 0.68 (0.67-0.69) |
| LDTEWS | 0.18 (0.17-0.18) | 0.52 (0.52-0.53) | 0.75 (0.74-0.76) | 0.52 (0.52-0.53) | 0.01 (0.01-0.01) | 0.62 (0.61-0.63) |
| LDTEWS:NEWS | 0.21 (0.20-0.21) | 0.67 (0.66-0.68) | 0.77 (0.76-0.79) | 0.67 (0.66-0.68) | 0.02 (0.02-0.02) | 0.80 (0.79-0.80) |

**Table 3:** The performance of machine learning models and the corresponding feature weights for the most and least important features; Sections A, B, C, and D explore the performance of the best machine learning model (GBT model on the F9 feature set). Section A outlines the performance of the accuracy optimised threshold for the GBT model. The threshold was set on the training data and tested on the test data. Section B outlines the performance of the GBT model after limiting the predictors to the 20 most important features. Sections C and D outline the performance of the GBT model after reducing the look back window from 24 hours to 6 and 12 hours. Section E outlines the performance after adding FiO2 as a predictor. FiO2 did not improve model performance compared to the optimised threshold model (section A of table 3); however, it ranked as the most important feature on feature importance analysis. Section F outlines the performance of the model after adding age as a predictor. Age did not rank within the most important features and did not improve performance over the performance of the optimised threshold model (section A of table 3). Section G outlines the performance after adding delta baseline to the vital signs and variations feature set. Section H outlines the performance after adding delta baseline to the all features and vital variations feature set. Section I outlines the performance of the GBT model with the hard output threshold on the different feature spaces before identifying the best model and adjusting the threshold for accuracy. Section J outlines the feature importance for the GBT model on the F9 feature set. It includes the nine most and least important features.

| Section | Feature | Model | Threshold<br>(on train) | Acc | Sen | Sps | Prs | AUROC |
| --- | --- | --- | --- | --- | --- | --- | --- | --- |
| A: Accuracy-optimised threshold | F9 | GBT | 0.12<br>(0.11-0.13) | 0.70<br>(0.69-0.71) | 0.96<br>(0.95-0.96) | 0.70<br>(0.69-0.70) | 0.03<br>(0.03-0.03) | 0.94<br>(0.94-0.94) |
| B: Feature selection | F9<br>(top 20) | GBT | 0.35<br>(0.32-0.37) | 0.80<br>(0.80-0.81) | 0.91<br>(0.90-0.92) | 0.80<br>(0.80-0.81) | 0.04<br>(0.04-0.04) | 0.94<br>(0.94-0.94) |
| C: 6-hour lookback window | F9 | GBT | 0.09<br>(0.08-0.09) | 0.56<br>(0.56-0.57) | 0.87<br>(0.86-0.88) | 0.56<br>(0.56-0.56) | 0.01<br>(0.01-0.01) | 0.85<br>(0.85-0.85) |
| D: 12-hour lookback window | F9 | GBT | 0.32<br>(0.30-0.34) | 0.66<br>(0.65-0.67) | 0.87<br>(0.86-0.88) | 0.66<br>(0.65-0.67) | 0.02<br>(0.02-0.02) | 0.86<br>(0.86-0.86) |
| E: Adding FiO2 as a predictor | F9<br>& FiO2 | GBT | 0.15<br>(0.13-0.18) | 0.72<br>(0.71-0.73) | 0.89<br>(0.87-0.91) | 0.72<br>(0.71-0.73) | 0.03<br>(0.03-0.03) | 0.93<br>(0.93-0.93) |
| F: Adding Age as a predictor | F9<br>& age | GBT | 0.19<br>(0.17-0.21) | 0.73<br>(0.72-0.74) | 0.94<br>(0.94-0.95) | 0.73<br>(0.72-0.74) | 0.03<br>(0.03-0.03) | 0.93<br>(0.93-0.94) |
| G: Adding Delta baseline to vital signs and delta | F10 | GBT | 0.32<br>(0.30-0.34) | 0.82<br>(0.81-0.83) | 0.87<br>(0.86-0.88) | 0.82<br>(0.81-0.83) | 0.04<br>(0.04-0.04) | 0.93<br>(0.92-0.93) |
| H: Adding Delta baseline to all features and delta | F11 | GBT | 0.22<br>(0.21-0.24) | 0.74<br>(0.73-0.74) | 0.93<br>(0.92-0.93) | 0.73<br>(0.73-0.74) | 0.03<br>(0.03-0.03) | 0.93<br>(0.92-0.93) |

| Hard output performance (Section I) |  |  |
| --- | --- | --- |
| Feature | Model | AUROC |
| F1 | GBT | 0.83 (0.83-0.84) |
| F2 | GBT | 0.72 (0.71-0.72) |
| F3 | GBT | 0.85 (0.84-0.85) |
| F4 | GBT | 0.77 (0.76-0.77) |
| F5 | GBT | 0.69 (0.68-0.69) |
| F6 | GBT | 0.78 (0.78-0.79) |
| F7 | GBT | 0.93 (0.92-0.93) |
| F8 | GBT | 0.86 (0.86-0.87) |
| F9 | GBT | 0.94 (0.94-0.94) |
| F10 | GBT | 0.93 (0.92-0.93) |
| F11 | GBT | 0.93 (0.93-0.93) |

| Feature weights (Section J) |  |  |  |
| --- | --- | --- | --- |
| Highest feature weights |  | Lowest feature weights |  |
| FiO2 | 0.258354 | Bilirubin-umol/L | 0.000031 |
| Max-Min SBP | 0.151461 | METHB (BG) | 0.000020 |
| CRP-mg/L | 0.108911 | FCOHB (BG) | 0.000011 |
| Max-Min HR | 0.044093 | CLAC (BG) | 0.000009 |
| PO2 (BG) | 0.033090 | NA+ (BG) | 0.000007 |
| Mean CellVol-fL | 0.026848 | Basophils-x10<sup>9</sup>/L | 0.000003 |
| CA+ + (BG) | 0.026313 | masktyp | 0 |
| Max-Min RR | 0.025169 | TEMPERATURE POCT | 0 |
| CTO2C (BG) | 0.024777 | Potassium-mmol/L | 0 |
| TEMP | 0.021500 | avpu | 0 |

We conducted three additional experiments. The first was to limit the predictors of the GBT model to the 20 features that ranked the highest on the feature importance scale considering the training set. This experiment did not impact performance (Table 3). The second experiment was to include a more granular measurement of oxygen support. We included the Fraction of Inspired Oxygen (FiO2) for this aim. Including the FiO2 did not improve the performance (Table 3). The third experiment was to include age as a predictor. Including age as a predictor did not significantly impact the performance (Table 3). The lack of performance gains in spite of the high feature importance may be due to multicolinearity, where a subset of existing variables highly correlate with this feature. This is explicit in the construction of the FiO2 variable, which is calculated from source variables already present in the Vital Signs feature set (Respiratory Rate, SpO2, Masktype) as outlined in the methods section.

Our results show that summary measures of variability of vital signs and laboratory markers play an important role in predicting deterioration. Adding the variability (range, mean of previous 24-hour window) and delta (current value - mean) features to the vital signs feature set added 10 percentage points to the AUROC (vital signs 83% vs vital signs & variations 93%). Similar results were observed in the all features feature set, where adding the variability and delta predictors added 8 percentage points to AUROC (all features 86% vs all features & variations 94%). Adding the delta baseline variables to both the all feature and vital signs feature spaces has improved the performance (vital signs & variations & baseline 93%; all features & variations & baseline 93%). These observations echo common clinical practice where physicians often analyse trends of parameters rather than their absolute values when evaluating a patient and highlight the benefits of dynamic monitoring. Moreover, the importance of summarising the variability and changes of vital signs when using them as inputs for machine learning models has already been demonstrated by Shamout and colleagues [31] in their work to develop a Deep Learning-based Early Warning System.

The lower performance of the model when using variables from blood gas analysis could partly be explained by inconsistency in the labelling of these samples. The origin of the blood, whether venous or arterial, was frequently missing or mislabelled perhaps reflecting time pressures on clinical staff, or skewed where interest is towards markers minimally influenced by sample provenance (e.g. lactate). This required the use of imputation techniques during the preprocessing of the dataset, which may have had an effect on performance. Moreover, some data points in blood gas readings duplicated information encoded within other feature sets, such as haemoglobin and creatinine.

### 2.3. Classification and misclassification

To assess for biases in model performance, we assessed rates of patient misclassification during validation for the best performing ML technique. We observed that rates of misclassification were higher for white (44%) than black, Asian and minority ethnic group patients (22%). The misclassification rate was similar between men (47%) and women (42%) and between patients aged over 60 (43%) and patients aged between 18 and 60 (44%).

## 3. Discussion

In this study, we assess the performance of existing EWS for predicting escalation to high-level oxygen support or unplanned ICU admission; this is an area of clinical importance in COVID-19. The EWS studied have been previously validated for predicting events such as cardiac arrest, unplanned ICU admission or death (Supplementary A: Table S6). However, limitations of using death as an outcome measure include that the score may be identifying an early sign of an already irreversible process, and therefore early identification of this may offer limited opportunity for clinical intervention. By contrast, our COVID-19 focused outcome measure provides a clinically useful and actionable warning which may help clinicians and healthcare system managers to preempt shortages and optimise resource allocation in a pandemic context. The difference in performance between our model and the EWS could be partially explained by the fact that these EWS were developed and optimised to detect ward patients’ deterioration against different outcomes. Nonetheless, these EWS represent the current standard of care for COVID-19 patients, and we took action to mitigate these effects by optimising each EWS threshold for our COVID-19 inpatient cohort.

A strength of our machine learning approach is its interpretability, using methods employed elsewhere in clinical practice [32] and shown able to attain patient and clinician trust. The three selected models (GBT, random forest and linear regression) permit querying of variables’ weights and presentation in an explainable way. This ability to make sense of the algorithm decision-making process has repeatedly been described as a critical factor in increasing technology uptake in clinical practice [33]. Moreover, our feature sets are oriented around routinely collected clinical data collected within existing care pathways, including calculation of EWS scores. Our models are therefore rapidly deployable within current clinical pathways.

A relative limitation of our study is that the number of features approximates that of patients within the training set. There is consequently a risk of overfitting when considering all the clinical features available, as exemplified by the increase in performance when limiting our inputs to the 20 most influential variables. Additionally, while a prediction window >1hr before an event is in line with existing vital monitoring systems [32], the window between sets of vital signs and positive events may capture overlapping transition effects. For example, an escalation in FiO2 may represent an emergency response to physiological deterioration which is necessarily followed by escalation of oxygen delivery device and ICU admission (where that level of support can be provided). Therefore, predictions made after a rapid escalation in FiO2 may capture patients where a clinical deterioration has already occurred, and ICU admission/higher level respiratory support is presently being arranged. Increasing the window upper bound to 3 hours before an escalation event had minimal effect on performance. On examination, the majority of escalation events (109/137) contained observations within a 12-24 hour prior window. Therefore, our data suggests that this is not a significant limitation in our case. The multivariate nature of EWS permit partial scores to be calculated where data is missing, however the machine learning methods examined require prior handling of missing data. These can be challenging to impute, as clinical data is often not missing at random. Missing data may therefore be poorly represented by population average values; for example, recording of vital signs is performed less frequently where there is no clinical concern. Limitations of imputation strategies include also the loss of important metadata. The presence of a measurement can often encode clinical meaning, for example the presence of an arterial blood gas reading is often driven by clinical concern of respiratory compromise; semantic knowledge which is lost by imputation. Nonetheless, in this study we demonstrate minimal difference in model performance across a range of imputation strategies on model performances, demonstrating minimal difference. By contrast, the multivariate, interpretable nature of EWS permit a partial score to be calculate despite missingness.

In this study, we assessed the theoretical performance of three machine learning approaches against some of the current EWS. Translation to clinical practice requires further optimisation and prospective valuation in a representative clinical population. Such optimisations include a better understanding of the dynamic evolution and availability of clinical data in real-time in the healthcare setting. Calibration of alarm trigger thresholds should be guided by desired clinical performance, reflecting healthcare system resource constraints and priorities, and a product design accounting for an optimal human-computer interaction.

## Data Availability

Access to data can be sought through applying to access our dataset via the formal application process.

## 4. Acknowledgement

This research was supported by the National Institute for Health Research (NIHR) Oxford Biomedical Research Centre (BRC). The views expressed are those of the authors and not necessarily those of the NHS, the NIHR or the Department of Health.

## Methods Section

### Data source

Deidentified data from patients were obtained from the Infections in Oxfordshire Research Database (IORD) which has Research Ethics Committee, Health Research Authority and Confidentiality Advisory Group approvals (19/SC/0403, ECC5-017(A)/2009). The dataset includes administrative data, vital-sign measurements, laboratory test results, and data on the level of oxygen support. We specifically extracted the data of patients who received a positive COVID-19 diagnosis between March 13^th^ and July 30^th^ 2020. 2,662 patients tested positive for COVID-19 and 612 of those patients were admitted within a window 48 hours prior to positive test to 30 days after. Only the admitted patients were included in the dataset and 101 patients who had a ‘Do Not Resuscitate’ status prior to their COVID test were excluded as their therapy may not have been escalated beyond ward-based care despite respiratory deterioration. Patients who were immediately escalated to advanced respiratory care upon admission (i.e. within 1 hour) were also excluded. The final dataset included 472 patients. Our model features were derived from four sets of commonly collected clinical variables: physiological variables, demographic information, oxygen support level, and laboratory test results (Table 1). The most recent previous blood tests within 5 days of the vital signs observations were considered.

### Feature Sets for EWS Systems

The EWS systems assessed each patient for deterioration every time vital signs were measured (NEWS, CEWS, MCEWS, AEWS) or when lab test results were obtained (LDTEWS and LDTEWS:NEWS). Oxygen support was used as a binary predictor for all of the EWS systems, except for LDTEWS and CEWS (in which it was not considered as a predictor in the original publication) [3, 9].

### Outcome definition

We defined the outcome of deterioration as either an escalation in the level of oxygen support requirements to either a level 2 or level 3 delivery device, or an unplanned ICU admission within a window of 24 hours. To do this, we created 4 levels of oxygen support based on the respiratory support device used (level 0: room air, level 1: low-flow oxygen support devices (flow less than 10L/Min, e.g. nasal cannulae), level 2: oxygen support devices with a flow over 10L/Min (e.g. reservoir bag), and level 3: high-flow ventilation or invasive ventilation). A detailed list of the oxygen support devices used to make the classification is available in the supplementary materials (Supplementary A: Table S5). We have defined progression to level 2 or 3 devices as an escalation to high-level oxygen support; therefore, a patient who progressed from L0 or L1 to L2/L3/unanticipated ICU admission would be considered to have deteriorated, while a transition from L0 to L1 would not be considered a deterioration. L2 indicates a deterioration in the condition of the patient and an increase in the need for respiratory support, while L3 is an advanced level of support that is dependent on respiratory support equipment that is in limited supply (i.e. ventilators or non-invasive ventilation equipment). Outside the scope of this paper, predicting L0 to L1 deterioration can be clinically valuable as it would differentiate patients who can be discharged (patients on room air who do not require oxygen support) from those who need hospital admission (requiring L1 or above support), and a trigger for starting dexamethasone therapy [34, 35]. However, we hypothesised that identifying the patients who need (L3) or are expected to need (L2) the advanced level of support would be more valuable because it provides the clinical teams the opportunity to optimise the management of resources that are in short supply during an event like a pandemic.

### Machine Learning models

We investigated the performance of (i) a basic machine learning classifier: Logistic Regression (LR) and (ii) two ensemble learning methods: Random Forest (RF) and Gradient Boosting Trees (GBT). Details of each method, parameter settings, and their strengths and weaknesses are shown in supplementary materials (Supplementary A: Table S8).

### Feature Sets for Machine Learning Models

To evaluate the EWS performance and compare it with that of the machine learning models, multiple feature sets were considered (Table 1). To ensure fairness, we first established a baseline comparison on the same feature sets as employed by each EWS. For example, the vital signs feature set is used by NEWS, CEWS, MCEWS, AEWS, and hence their performance was compared with an ML model using the same input features. Similarly, for the EWS bloods feature set (used by LDTEWS) and the vital signs & EWS bloods feature set (LDTEWS:NEWS), the same inputs were considered for the comparative ML models. In addition, we trained and evaluated the ML models on various other feature sets. Six sets of clinical parameters were investigated: (1) 24 routinely collected laboratory blood tests, (2) 21 routinely measured/estimated point-of-care blood gas readings, (3) changes in vital signs results in a window of 24 hours before the given observation, (4) measurements of seven routinely measured physiological parameters, (5) variance of the current vital signs from a baseline of a previous admission, and (6) variance of the current vital signs, blood tests, and blood gases from a baseline of a previous admission. The components of each feature set are detailed in Table 1. Pre-existing oxygen support before the point of prediction was indicated by a binary variable (1 for L1 support and 0 for L0 support) (Supplementary A: Table S5). Consequently, we considered the following feature sets for ML analysis: (vital signs – F1) vital signs; (EWS bloods – F2) EWS bloods feature set; (EWS blood & vital signs – F3) a combination of F1 and F2 feature sets; (blood tests – F4) clinical parameters in (1); (blood Gas – F5) clinical markers in (2); (blood tests and gas – F6) a combination of F4 and F5 feature sets; (vital signs & delta – F7) a combined feature set of F1 feature set and (3); (all features – F8) a combination of F1, F4, and F5; (all features & delta – F9) a combined feature set of F3, F4, F6; (vital signs & delta baseline & delta – F10) a combination of F1, (3), and (5); and (all features & delta baseline & delta – F11) a combination of F3, F4, F6, and (6).

### Calculation of the FiO2 values

Fraction of inspired oxygen (FiO2) values (%) were calculated based on the mask type used. Depending on the mask type, oxygen flow (O2 flow, L/min) and patient’s respiratory rate (RR, breaths/min) were included in the calculation. Simple face masks, nebuliser masks, tracheostomy masks and Oxy-Masks were considered as Hudson masks.

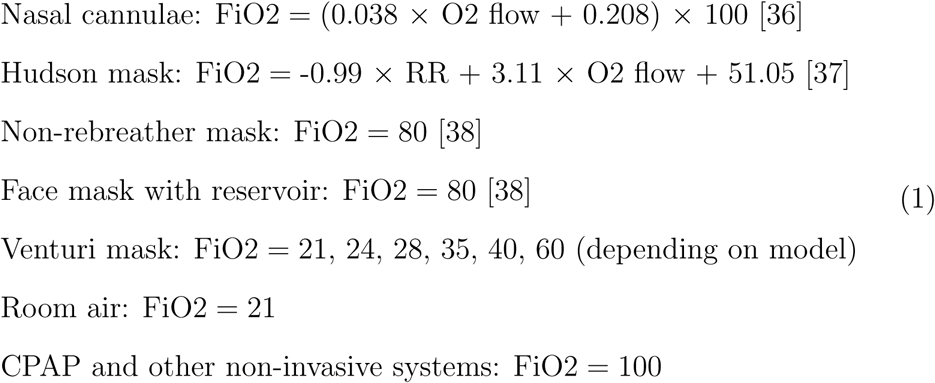

### Data preprocessing

We treated observation sets as independent rather than as grouped by patient admission. We excluded implausible physiological values. Non-numerical readings were replaced with clinically appropriate values. Where a lab value was reported as being below the threshold of detection of the laboratory assay, the value was replaced with a numerical zero value. Where values were reported as being above the threshold of detection, clinically appropriate values were selected to maintain the significance of the high result. When the provision or absence of supplemental oxygen was missing, we assumed that supplemental oxygen was not provided and set the supplemental oxygen value to 0. Similarly, we have made the same assumption for AVPU and replaced missing AVPU values by ‘alert’. For missing values, we have used multiple imputation techniques (mean, median, Bayesian ridge regression, and stochastic regression) to compensate for missing values across the dataset. The best performing imputation method across different experiments was median, hence we have chosen it as the default method in our analysis as a design choice (Supplementary A: Figure S1).

### Alerting Thresholds

The evaluated EWS systems (Table 2) are provided with default alerting thresholds to convert the computed score to ‘alert’ or ‘no alert’. NEWS gives an individual score of 2 when a patient is on supplementary oxygen, and an individual score of 0 when the patient is on room air. NEWS aggregates the individual scores to an overall score which is assessed against an alerting threshold (default 5). We used the default individual scores for EWS. However, given that we are predicting an outcome different from the default predicted outcome of the EWS (escalation in oxygen demand vs in-hospital mortality, cardiac arrest or unplanned ICU admission), we chose to evaluate the performance of the EWS not only based on the original overall thresholds (e.g. 5 for NEWS) but also on optimised thresholds.

We optimised the machine learning and EWS thresholds to report the performance metrics on the test set by identifying the thresholds that maximise the accuracy on the train dataset, and used these thresholds on the test set.

### Performance assessment

For all experiments, the classification was performed by training on a balanced dataset and then testing on an imbalanced (representative) dataset. We ran the classification over multiple iterations and cross fold validations. In each fold, 20% of the data was considered as the test set. Within the remaining 80% of the data, since non-events outnumbered events, non-events were sub-sampled randomly to balance the size of the two classes. This was run over 40 iterations of 5-fold stratified cross-validation. We chose k-fold stratified due to the imbalanced nature of the classes.

For machine learning, Gradient Boosting Trees, Random Forest, and logistic regression were considered as basic machine learning techniques (Supplementary A: Table S8). For EWS, the test set was used to calculate EWS scores in each fold.

We evaluated the performance of our EWS and machine learning methods using the AUROC to predict an outcome of deterioration defined as either escalation in the level of oxygen demand (to level 2 or 3) (Supplementary A: Table S5) or an unplanned ICU admission. The performance in terms of accuracy, sensitivity, specificity, precision, and AUROC were calculated for the validation sets (for parameter setting) and test sets (for final comparison) and averaged over iterations; mean and standard deviation were reported.

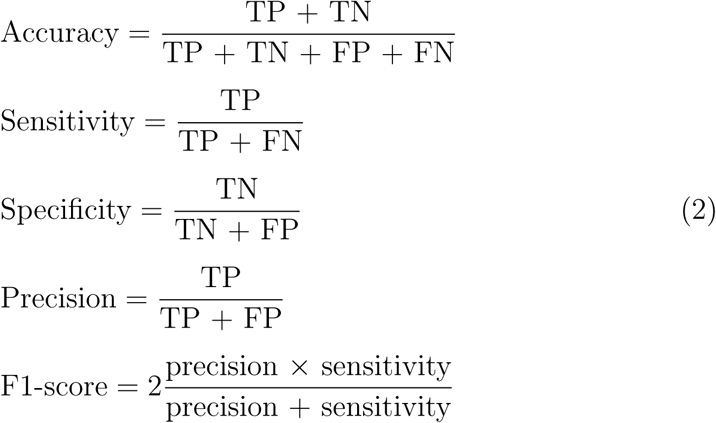

TP, TN, FP, and FN stand for true positive, true negative, false positive, and false negative, respectively. Considering a probability estimate as the output of each classifier for the validation set and setting various thresholds to categorise this output as event/non-event could result in different TP, FP, FN, and TN rates. Alternatively, a receiver operating characteristic (ROC) curve showing the sensitivity as a function of 1 - specificity for different thresholds; each point in the curve indicates a specific value for sensitivity, specificity, and accuracy. AUROC-ROC is the area under the ROC curve. The parameters of the models (e.g., number of ensembles for RF or GBT) were optimised through the internal cross-validation on the training data. This was done by a grid search over a range of values and selecting parameters that generated the best AUROC-ROC. The model with the highest performance was reported in the paper.

### Patient and public involvement

The IORD panel, which includes patient and public representatives provided feedback on the study design and approved its final form.

**Figure 1:**
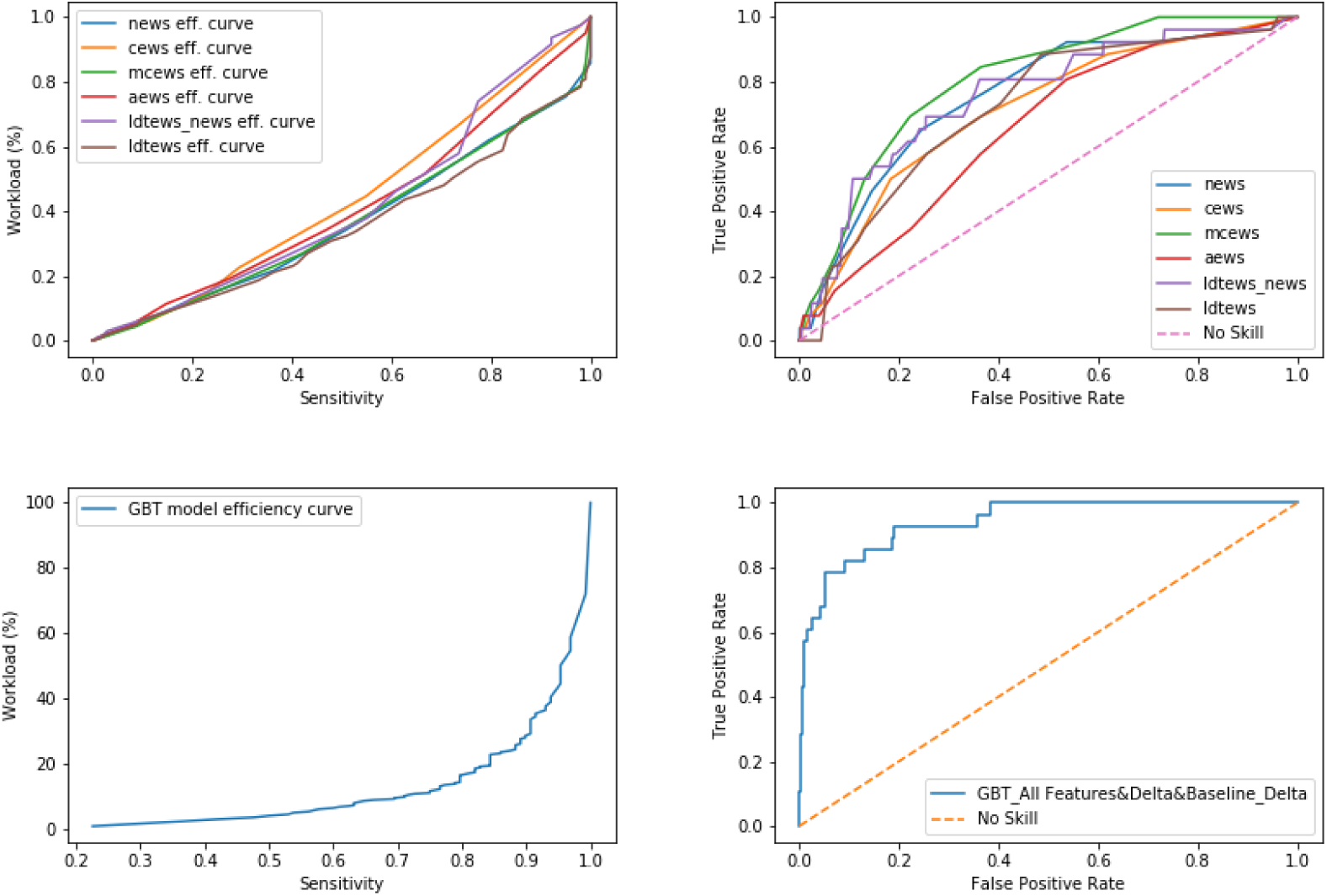
This figure includes the efficiency and Receiver Operating Characteristic (ROC) curves for the machine learning models and the Early Warning Scores (EWS). The upper right figure illustrates the ROC curves for the various EWS in our study (the best performance is for NEWS with AUROC of 72%). The upper left figure illustrates the efficiency curves for the EWS in our study. The low performance of the EWS on the Efficiency Curve metric may be explained by a high false positive. The lower right figure illustrates the ROC and AUROC for the GBT model on F11 feature set (AUROC of 90%). The lower left figure illustrates the performance of the GBT model measured by the efficiency curve metric.

