## Supplementary materials for "Development and Validation of Early Warning Score Systems for COVID-19 Patients"

Table 1: The descriptive statistics of the study dataset

| **Variable** | **Value** |
|---|---|
| Age (mean±std) | 67.85±16.45 |
| Age >45 (%) | 91.11% |
| Female (%) | 46.99% |
| Admissions ($n$) | 472 |
| Events ($n$) | 137 |

Table 2: Mean and IQR of Vital Signs in the study dataset

| **Vital Sign** | **Mean (IQR)** |
|---|---|
| Heart Rate (bpm) | 89.7 (77.0-100.0) |
| Respiratory Rate (brpm) | 21.4 (18.0-24.0) |
| Systolic Blood Pressure (mmHg) | 127 (113-140) |
| $SpO_2$ (%) | 93.9 (93.0-96.0) |
| Temperature (°C) | 37.1 (36.5-37.7) |

Table 3: Mean and IQR of Blood Tests in the study dataset

| **Test** | **Mean (IQR)** |
|---|---|
| Alanine Aminotransferase (IU/L) | 33.6 (18.0-40.0) |
| Albumin (g/L) | 27.8 (24.0-31.0) |
| Alkaline Phosphatase (IU/L) | 97.4 (62.0-111.5) |
| Basophils (x$10^9$/L) | 0.02 (0.01-0.02) |
| Bilirubin ($\mu$mol/L) | 10.1 (6.0-12.0) |
| C-reactive Protein (mg/L) | 148.8 (84.9-210.8) |
| Creatinine ($\mu$mol/L) | 148.5 (65.0-108.0) |
| Eosinophils (x$10^9$/L) | 0.03 (0.00-0.03) |
| Haematocrit (L/L) | 0.36 (0.32-0.43) |
| Haemoglobin (g/L) | 119.4 (102.0-141.0) |
| Lymphocytes (x$10^9$/L) | 1.0 (0.4-1.0) |
| Mean Cellular Volume (fL) | 90.4 (86.7-94.6) |
| Monocytes (x$10^9$/L) | 0.47 (0.22-0.57) |
| Neutrophils (x$10^9$/L) | 6.21 (4.02-8.01) |
| Platelets (x$10^9$/L) | 199.2 (140.0-230.5) |
| Potassium (mmol/L) | 4.0 (3.7-4.2) |
| Sodium (mmol/L) | 134.7 (132.0-138.0) |
| Urea (mmol/L) | 8.9 (4.5-9.3) |
| White Blood Cells (x$10^9$/L) | 7.8 (4.9-9.4) |

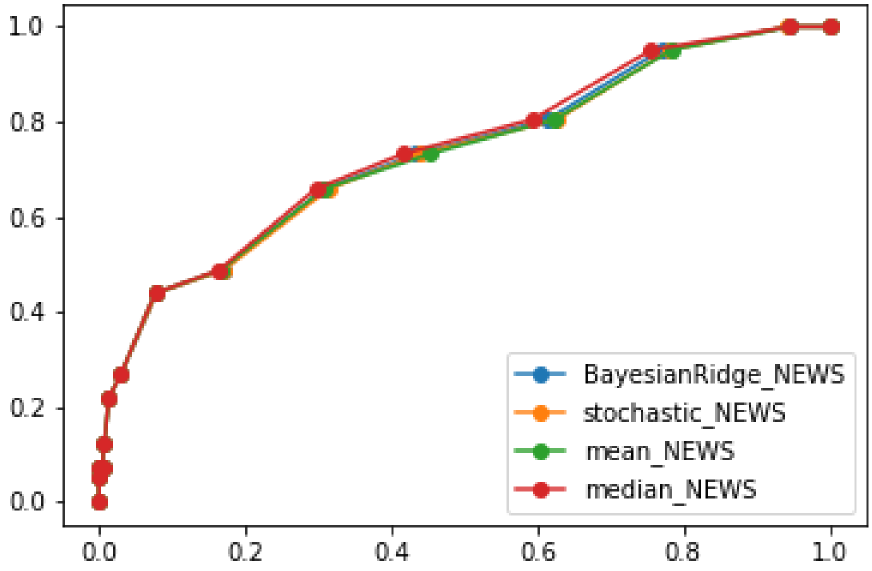


Figure 1: Performance of different imputation techniques for the NEWS score (As measured by the Area Under Receiver Operating Curve). The median technique has slightly outperformed the other imputation techniques. We have used the median as the imputation technique in our study. The x-axis is the False Positive Rate and the y-axis is the True Positive Rate

Table 4: Mean and IQR of Blood Gases in the study dataset

| **Test** | **Mean (IQR)** |
|---|---|
| BE ACT | 0.99 (-0.80-3.3) |
| BE STD | 1.05 (-1.10-3.60) |
| Bicarbonate | 24.9 (22.8-26.7) |
| Calcium | 1.10 (1.06-1.15) |
| cLac | 1.6 (1.0-1.8) |
| Creatinine | 170.1 (77.2-128.0) |
| $CtO_2C$ | 12.3 (8.0-16.7) |
| Est. Osmolality | 278.9 (271.6-284.6) |
| Fraction COHb | 0.95 (0.70-1.10) |
| Fraction HHb | 27.78 (6.70-47.70) |
| $FIO_2$ | 24.2 (21.0-21.0) |
| Glucose | 8.0 (5.8-8.7) |
| Haemoglobin | 125.5 (109.0-146.0) |
| Haematocrit | 38.5 (33.7-44.7) |
| Potassium | 4.0 (3.7-4.2) |
| Met. Hemoglobin | 0.7 (0.4-0.8) |
| Sodium | 135.2 (132.0-138.0) |
| $O_2$ Saturation | 71.0 (49.4-93.2) |
| P5OC | 3.6 (3.3-3.9) |
| $pCO_2$ | 5.15 (4.52-5.72) |
| pH | 7.43 (7.39-7.47) |
| $pO_2$ | 7.37 (3.94-8.74) |
| Temperature | 37.1 (37.0-37.0) |

Table 5: Different levels of O2 support and the corresponding oxygen support devices in each category.

| Level of O2 Support | Device Name |
|---|---|
| Level 0 | Room air (No Device support) |
| Level 1 | Nasal Cannulae, Simple Mask, Venturi Face Mask, Nebuliser Mask, Oxy-mask, Tracheostomy Mask |
| Level 2 | Non-rebreather mask, Reservoir Mask |
| Level 3 | High flow, Non-invasive system, CPAP, Airvo |

Table 6: Various Early Warning Scores evaluated in our study.

| Early Warning Score | Summary | Input data | Predicted outcome |
|---|---|---|---|
| National Early Warning Score (NEWS) [2] | The national EWS in the UK and one of the most widely used EWS | HR, RR, Supplemental O2, SpO2, SBP, Temperature, Level of Consciousness (AVPU) | Early Cardiac Arrest, Unanticipated ICU Admission, and in-hospital mortality |
| Centile-based Early Warning Score (CEWS) [1] | A centile-based early warning score using continuously acquired bedside vital-sign data | HR, RR, SpO2, SBP, Temperature, Level of Consciousness (AVPU) | Cardiac Arrest, Unanticipated ICU Admission, and in-hospital mortality |
| Manual Centile-based Early Warning Scores (MCEWS) [5] | A centile-based early warning score using manually-recorded data | HR, RR, Supplemental O2, SpO2, SBP, Temperature, Level of Consciousness (AVPU) | Cardiac Arrest, Unanticipated ICU Admission, and in-hospital mortality |
| Age-based Early Warning Score (AEWS) [6] | Age specific early warning score based on the NEWS score | HR, RR, Supplemental O2, SpO2, SBP, Temperature, Level of Consciousness (AVPU) | Cardiac Arrest, Unanticipated ICU Admission, and in-hospital mortality |
| Laboratory Decision Tree Early Warning Score (LDTEWS) [3] | An early warning score (EWS) based on routinely collected laboratory tests | HGB, Alb, Na+, K+, Cr, Ur, WBC | In-hospital mortality |
| LDTEWS: NEWS [4] | An EWS developed by combining (NEWS and LDTEWS) to discriminate unanticipated ICU admission | NEWS and LDTEWS input data | In-hospital mortality and unplanned ICU admission |

Table 7: Classification methods, their parameter settings, and pros and cons.

| Method | Summary | Parameter setting | Pros and cons |
|---|---|---|---|
| LR | LR is a linear model that optimises a set of weights for each feature that lead to the best classification performance. | This model was performed using LIBLINEAR library. | LR is easy to implement and efficient to train. However LR cannot solve non-linear problems and has high bias. |
| RF | RF is an averaging method that is based on building several independent Decision Tree (DT) classifiers on different subsets of the dataset and average results to produce final predictions and improve the performance. | 100 estimators were considered for RF training. | It is based on training of several independent trees that can be fit in parallel. Moreover, RF usually reduce the variance. However, it needs heavier computational resources. |
| GBT | GBT is a boosting method based on sequentially building weak estimators (models that are only slightly better than the random prediction, i.e. small DTs). Initially, all data samples have the same weight. However, at successive iterations, the weights of mislabelled training samples by the boosted model at the previous step are increased, while the weights are decreased for the remaining training samples. Hence, as iterations proceed, the influence of difficult samples is increased. Later, it combines the predictions through a weighted majority vote with the aim of reducing the bias of the combined classifier. | DT is used as the base classifier and Binomial deviance and 100 estimators were considered for the training. | It is more robust to outliers and has high predictive power. Nonetheless, due to the sequential nature, it cannot be parallelised. |
